# POPULATION SCREENING FOR HUMAN PAPILLOMAVIRUS IN WOMEN IN THE METROPOLITAN AREA OF TUNJA CITY, BOYACÁ

**DOI:** 10.1101/2025.09.25.25336668

**Authors:** Lorenzo Hernando Salamanca Neita, Elizabeth Guío Mahecha, Mónica Gabriela Huertas Valero, Johana Marín Suárez, Juan Pablo Carvajal Rojas, Gloria Eugenia Camargo Villalba, Laura Ximena Ramírez López

**Affiliations:** Carvajal Laboratorios IPS S.A.S; Universidad de Boyacá

**Keywords:** Human Papillomavirus, detection, urine, cervicovaginal samples, concordance, sensitivity, specificity

## Abstract

**Background:** Human Papillomavirus (HPV) detection is traditionally performed using cervicovaginal or vaginal cuff samples. However, first-void urine has also proven to be suitable for detecting HPV DNA. This could facilitate access to viral screening tests. The objective of this research was to determine and compare the presence of HPV genotypes in cervicovaginal and urine samples from women in Tunja, Colombia.

**Methods:** This was a cross-sectional study conducted on 161 women aged 20 to 65 living in Tunja. Cervicovaginal/vaginal cuff and urine samples were collected to detect HPV genotypes. Concordance, sensitivity, specificity, and association with sociodemographic/gynecological variables were evaluated.

**Results:** HPV prevalence was 29.2% in cervicovaginal samples, while in urine it was 32.9%. Genotypes 52 and 68 were the most prevalent in both cervical and urine samples. Comparison of HPV detection between samples revealed weak concordance (kappa=0.392). Sensitivity and specificity of viral detection in urine were 61.7 and 78.9, respectively. A significant positive association was found in women aged 18-37 who had initiated sexual activity early and had menarche before age 13.

**Conclusions:** The prevalence of HPV infection was low. High-risk genotypes were identified in both types of samples. The diagnostic performance of the urine test was low, demonstrated by the heterogeneity of the results between the two samples.

## 1. INTRODUCTION

Human Papillomavirus (HPV) is one of the main causative agents of sexually transmitted diseases worldwide, infecting up to 80% of sexually active women at some point in their lives. Of these, 10-20% develop persistent infections [1]. Certain HPV strains have been associated with the development of Cervical Cancer (CC), a preventable and treatable malignancy with early detection through a Pap smear of precancerous Cervical Intraepithelial Neoplasia (CIN) [2]. However, this invasive detection technique requires specialized personnel and shows a concerning trend of low coverage in women under 35 years old [3], possibly due to its invasive nature, time consumption, and the need for trained personnel to collect samples.

Currently, the HPV DNA detection test in cervical cells is the reference method for identifying infection [4]. However, in recent years, there has been growing interest in the use of non-invasive self-sampling methods, such as first-void urine. Numerous studies have demonstrated that urine is an adequate sample for detecting HPV DNA, with stability maintained using preservatives [5]. Additionally, it has been concluded that first-void urine sampling is a preferred non-invasive method, ensuring good concordance in viral DNA detection compared to reference cervical samples [6].

Given the above, the objective of this study was to determine and compare the presence of Human Papillomavirus genotypes in cervicovaginal and urine samples from women in the metropolitan area of Tunja, Colombia.

## 2. MATERIALS AND METHODS

An observational, cross-sectional analytical study was conducted on 161 women aged 20 to 65 living in Tunja, Boyacá, Colombia. The sample size was calculated considering a 10% loss, an estimated HPV infection prevalence of 10.6% [7], and a 95% confidence limit. The study was approved by the Ethics and Bioethics Committee of Universidad de Boyacá (memorandum RECT-147/2022).

Participants who had initiated sexual activity were included, and those with a prior CC diagnosis were excluded. After the awareness process, written informed consent was obtained from participants who completed a structured survey to collect sociodemographic and gynecological data. Samples were collected from September 13 to November 3, 2022.

### 2.1. Sample Collection

Each participant received a Colli-Pee device and was instructed to collect the first-void urine sample of the day without prior genital cleansing. Cervicovaginal samples were then taken using a cytobrush. This procedure was performed by a specialist gynecologist. In the case of previous hysterectomy, a vaginal cuff sample was taken.

### 2.2. Sample Processing

Urine samples were pre-treated by centrifugation, supernatant discard, and pellet resuspension in molecular-grade water, then stored at –80°C until processing. Cervicovaginal samples were stored at –80°C for nucleic acid extraction using the semi-automated Nextractor system (Genolution, Seoul, Korea) and the NX-48S kit, following the manufacturer’s protocol. Genetic material from urine and cervicovaginal samples was processed simultaneously using the INNO-LiPA HPV Genotyping Extra II kit (Fujirebio, Ghent, Belgium). This kit consists of two phases: First phase: Amplification of viral DNA by Polymerase Chain Reaction (PCR) using the INNO-LiPA HPV Genotyping Extra II AMP kit. This kit is designed to amplify a region of approximately 65 base pairs within the L1 region of the HPV genome using a consensus primer system. The obtained amplicons were stored at –80°C for subsequent hybridization.

Second phase: Reverse line hybridization (LiPA) and genotype identification using the INNO-LiPA HPV Genotyping Extra II kit. Amplicons obtained in the first phase were hybridized with specific oligonucleotide probes for 32 HPV genotypes, immobilized in parallel lines on a nitrocellulose membrane. Subsequently, incubation with detection and chromogenic reagents was performed to visualize lines corresponding to genotypes present in each sample.

Results were interpreted as positive or negative for HPV presence. Positive results were classified into high-risk genotypes (HPV 16, 18, 31, 33, 35, 39, 45, 51, 52, 56, 58, 59, and 68), probable high-risk (HPV 26, 53, 66, 70, 73, and 82), and low or unknown risk (HPV 6, 11, 40, 42, 43, 44, 54, 61, 62, 67, 81, 83, and 84), according to reactive lines visualized on the membrane.

### 2.3. Statistical Analysis

Sensitivity and specificity values for HPV detection in urine samples were determined. To identify circulating HPV genotypes in the studied population, participant results were organized according to sample type, and HPV presence was classified qualitatively (positive or negative).

With qualitative results, the concordance of tests in cervicovaginal and urine samples was calculated using Cohen’s kappa index with a 95% confidence interval (CI), considering Landis’ classification [8] for interpreting indices: <0.00 minimal; >0.00-0.20 insignificant; 0.21-0.40 weak; >0.41-0.60 moderate; 0.61-0.80 substantial; 0.81-1.00 almost perfect.

Data from surveys were systematized in Microsoft Excel and analyzed with IBM SPSS Statistics 28. Hypothesis tests were conducted with a significance level of p<0.05. Categorical sociodemographic variables were described using frequencies and proportions, while means and standard deviations were calculated for continuous variables. To evaluate the association between cervicovaginal test results and variables of interest, odds ratios (OR) with 95% confidence intervals (CI) were estimated, adjusting ORs through logistic regression. Additionally, sensitivity and specificity values of viral detection using urine samples were determined.

## 3. RESULTS

### 3.1. Sociodemographic Characteristics of the Population

The study included 161 participants, all with complete questionnaire information and high-quality cervicovaginal and urine samples for processing and analysis.

The participants’ ages ranged from 18 to 65 years, with an average age of 37.18 years and a standard deviation of 11.6. The age distribution, according to Resolution 3280 of 2018 [9], which considers life stages, was as follows: youth (18 to 28 years), 48 women (29.8%); adulthood (29 to 59 years), 111 women (68.9%); and old age (60 years and older), 2 participants (1.2%).

**Table 1.**
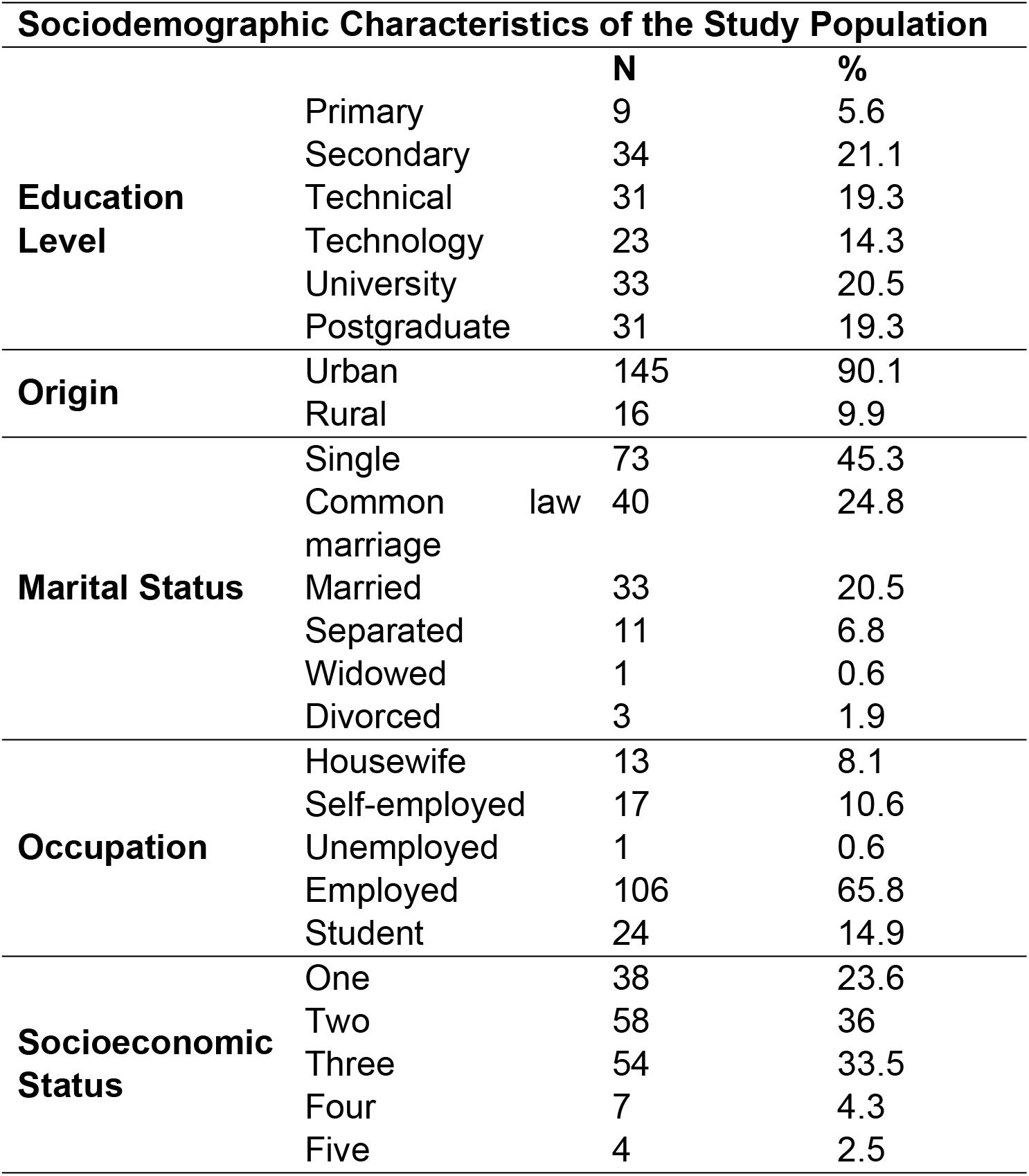
Sociodemographic Characteristics of the Study Population.

### 3.2. Gynecological History

Menarche occurred between the ages of 10 and 18, with an average age of 13.1 years. The age at first sexual intercourse ranged from 12 to 31 years, with a mean of 18 years. Thirty-eight women (23.6%) reported having no children, while the rest indicated having between 1 and 5 children, with one child being the most common response (27.3%). Over the past year, 70.8% of the participants (114 women) reported having had only one sexual partner. Regarding sexual practices, 28.6% (46 women) reported having oral sex, and 7.5% (12 women) reported having anal sex.

When asked about family planning methods, 16.1% (26 women) used barrier methods such as condoms, while 83.9% used other contraceptive methods, including intrauterine devices, oral contraception, monthly or quarterly injectable contraception, subdermal implants, transdermal patches, and emergency contraception. Additionally, 21.7% (35 women) reported having undergone tubal ligation.

Regarding Pap smear, 93.8% of the participants (151 women) had undergone this test at some point in their lives, and of these, 63.3% (102 women) had done so in the past year or currently. Moreover, 83.9% of the participants reported that their Pap smear results were normal.

### 3.3. HPV Detection and Genotype Distribution

The overall prevalence of HPV was 29.2% (47 women) in cervicovaginal samples, while the prevalence of HPV in urine samples was 32.9% (53 women). Eighteen participants had positive results in cervicovaginal samples but negative in urine samples for HPV. Conversely, 24 women had positive results in urine samples but negative in cervicovaginal samples. Additionally, 29 participants showed positivity in both cervicovaginal and urine samples.

Genotypes 52 and 68 were the most prevalent in both the cervix and urine samples. Genotypes 16, 45, 51, 52, 54, 58, and 66 were most prevalent only in cervicovaginal samples, while genotypes 51, 52, 53, 59, and 61 predominated in urine samples. There was a notable presence of high-risk genotypes in both types of samples analyzed.

**Table 1.** Distribution of HPV Genotypes by Sample Type (Cervicovaginal and Urine)

| Risk Type | Genotype | Both Positive | Only Swab | Only Urine | Both Negative |
| --- | --- | --- | --- | --- | --- |
| High Risk | HPV16 | 1 | 2 | 1 | 157 |
|  | HPV18 | 1 | 1 | 1 | 158 |
|  | HPV31 | 0 | 0 | 2 | 159 |
|  | HPV33 | 0 | 0 | 1 | 160 |
|  | HPV35 | 1 | 0 | 1 | 159 |
|  | HPV39 | 0 | 1 | 1 | 159 |
|  | HPV45 | 0 | 2 | 1 | 158 |
|  | HPV51 | 3 | 2 | 7 | 149 |
|  | HPV52 | 5 | 2 | 4 | 150 |
|  | HPV56 | 2 | 0 | 1 | 158 |
|  | HPV58 | 2 | 2 | 2 | 155 |
|  | HPV59 | 2 | 0 | 3 | 156 |
|  | HPV68 | 3 | 1 | 1 | 156 |
| Probable<br>High Risk | HPV26 | 1 | 0 | 0 | 160 |
|  | HPV53 | 2 | 1 | 3 | 155 |
|  | HPV66 | 2 | 2 | 2 | 155 |
|  | HPV70 | 2 | 0 | 2 | 157 |
|  | HPV73 | 0 | 0 | 0 | 161 |
|  | HPV82 | 1 | 0 | 2 | 158 |
| Low Risk | HPV6 | 0 | 0 | 1 | 160 |
|  | HPV11 | 0 | 0 | 0 | 161 |
|  | HPV40 | 0 | 0 | 0 | 161 |
|  | HPV42 | 0 | 0 | 0 | 161 |
|  | HPV43 | 0 | 0 | 0 | 161 |
|  | HPV44 | 2 | 1 | 1 | 157 |
|  | HPV54 | 0 | 2 | 1 | 158 |
|  | HPV61 | 0 | 0 | 6 | 155 |
|  | HPV62 | 1 | 0 | 0 | 160 |
|  | HPV67 | 2 | 0 | 0 | 159 |
|  | HPV81 | 0 | 1 | 1 | 159 |
|  | HPV83 | 0 | 0 | 1 | 160 |
|  | HPV89 | 0 | 1 | 2 | 158 |

Among the 47 women who tested positive for HPV in cervicovaginal samples, 7 exhibited the coexistence of two different genotypes, while 40 women presented only one genotype in these samples. Additionally, in the 53 women who tested positive in urine samples, it was found that 2 harbored four different genotypes, 4 women showed three genotypes, 14 women presented two genotypes, and 33 participants harbored only one genotype in urine samples.

**Table 2.** Distribution of Genotypes by Risk Between Urine and Cervicovaginal Samples.

| Genotypes |  |  |
| --- | --- | --- |
| Risk Type | Cervicovaginal | Urine |

|  | Samples |  | Samples |  |
| --- | --- | --- | --- | --- |
|  | N | % | N | % |
| High Risk | 27 | 57.4 | 24 | 45.3 |
| Probable High Risk | 9 | 19.1 | 9 | 17.0 |
| High and Probable High Risk | 1 | 2.1 | 5 | 9.4 |
| High and Low or Unknown Risk | 0 | 0 | 8 | 15.1 |
| High, Probable High, and Low or Unknown Risk | 0 | 0 | 1 | 1.9 |
| Low or Unknown Risk | 10 | 21.3 | 6 | 11.3 |

### 3.4 Concordance and Test Characteristics

**Table 3.** Test Performance Characteristics.

| Performance Characteristics of HPV Test in Urine |  |
| --- | --- |
|  | Result (95% CI) |
| **Prevalence** | **32.9** |
| Sensitivity | 61.7 (44.0-79.3) |
| Specificity | 78.9 (70.4-87.3) |
| Positive Predictive Value | 54.7 (36.5-72.8) |
| Negative Predictive Value | 83.3 (75.5-91.0) |
| Positive Likelihood Ratio | 2.92 (-3.20-9.04) |
| Negative Likelihood Ratio | 0.48 (-0.94-1.90) |

The comparison of molecular HPV detection between cervicovaginal and urine samples revealed a weak concordance, indicated by a kappa index of 0.392. For a more detailed evaluation of the characteristics of the urine test compared to the cervicovaginal test, the latter was considered the gold standard or reference test.

### 3.5. Factors Associated with HPV Presence

Results from cervical samples indicate a significant positive association in women aged 18 to 37 years (OR=2.7; 95% CI=1.3-5.6), in women who began sexual activity at age 15 or earlier (OR=3.2; 95% CI=1.3-7.9), and in those who experienced menarche before age 13 (OR=2.1; 95% CI=1.0-4.1). However, no statistically significant associations were observed with the other variables evaluated.

**Table 5.**
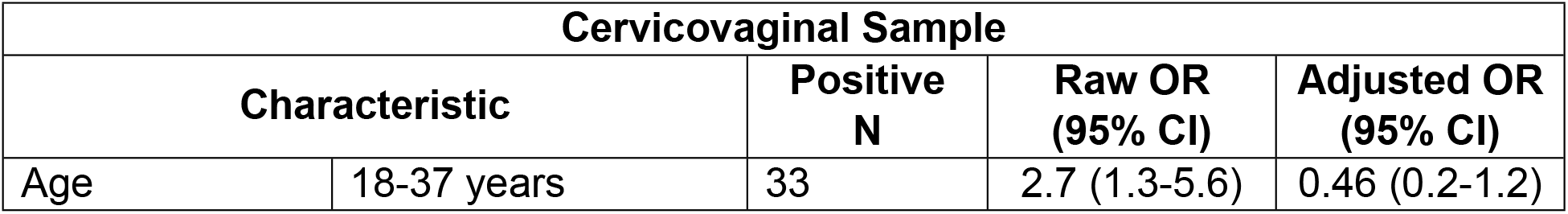
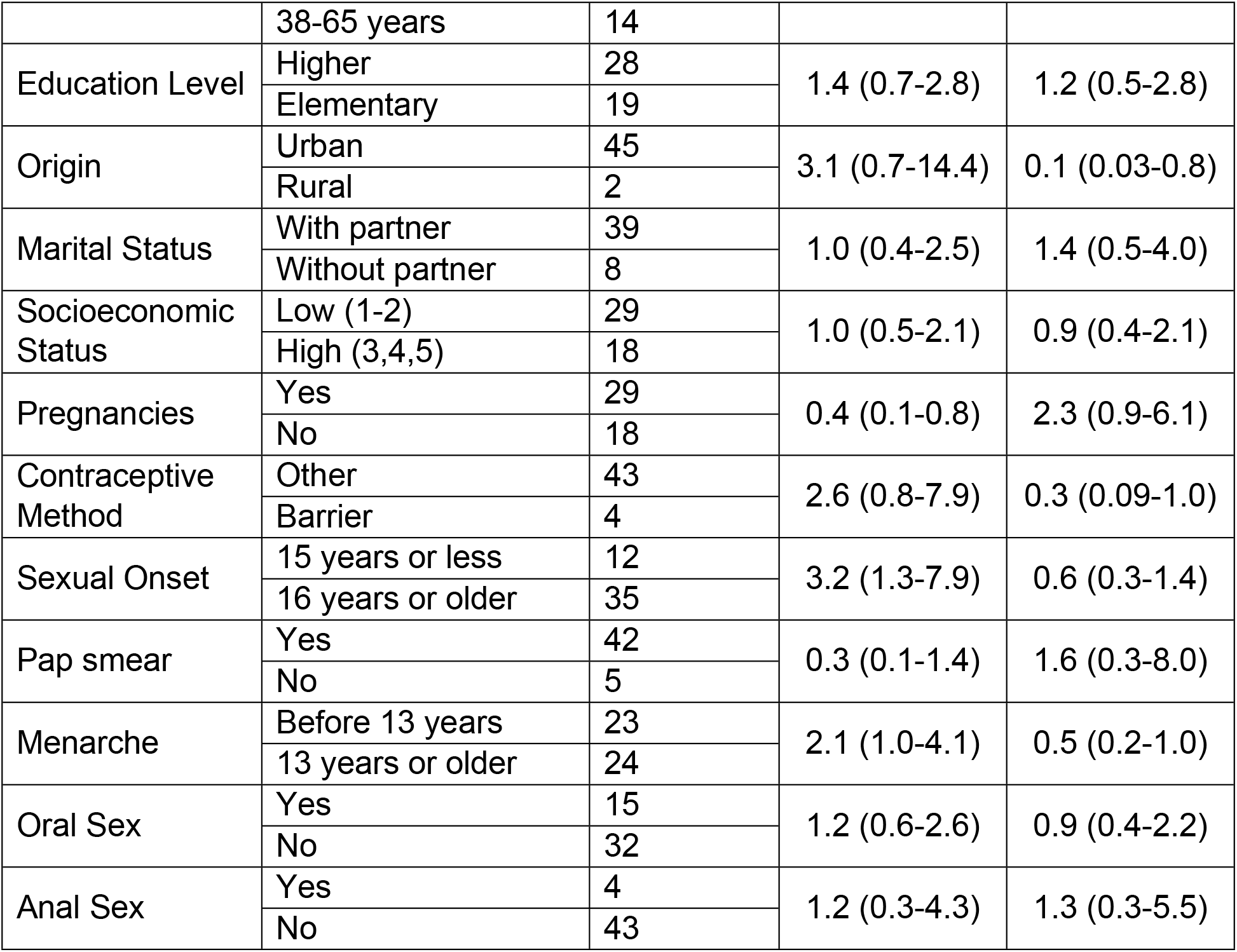
Factors Associated with HPV Presence.

## 4. DISCUSSION

HPV is one of the most common causes of sexually transmitted diseases worldwide [1]. Although most HPV infections are transient and asymptomatic, some may persist and progress to precancerous lesions or cervical cancer, especially those caused by high-risk genotypes. In Colombia, genital HPV infection is not a mandatory reportable disease, so the true incidence and prevalence figures by region are unknown [10].

HPV infection and cervical cancer represent a serious public health problem, especially in low- and middle-income countries like Colombia, where socioeconomic and educational factors have a significant impact, being the leading cause of years of life lost, devastating not only the women themselves but also their families [11]. In this study, most participants were from lower-middle socioeconomic status (2 and 3) and had higher educational levels, highlighting the importance of education in preventing this infection.

Both men and women are part of the HPV epidemiological chain and can be both carriers and victims of the infection [12]. Although this study did not show significant differences, factors associated with HPV infection reported in various studies include the number of sexual partners over a lifetime, sexual relationships with individuals at high risk for sexually transmitted infections, not using condoms in every sexual encounter, and oral sex practices [13-15].

Age is a determining factor in HPV infection prevalence. A 2017 study estimated that more than half of the high-risk HPV infections that progress to cancer are acquired by age 21 [16], while a meta-analysis reported that in all included studies, HPV infection prevalence decreased with increasing age, with the highest prevalence in younger women (≤25 years) [17]. Although most sexually active adults will be infected with HPV at least once in their lifetime, sexually active women under 25 consistently have the highest infection rates [18]. However, in our study, ages between 18 and 37 years were associated with a higher risk of HPV infection in cervicovaginal samples (OR=2.7; 95% CI=1.3-5.6). Additionally, early onset of sexual activity (before 14-16 years) also showed a significant association (OR=3.2; 95% CI=1.3-7.9), consistent with other studies [15,19].

It has been reported that the use of condoms and contraceptive injections over a lifetime is associated with a higher risk of HPV infection [20]. In this research, none of the gynecological histories studied were associated with HPV infection. The literature also describes a correlation between cervical cancer and the presence of HPV with co-infections by other agents in the vaginal tract, such as Herpes simplex type 2 and the bacteria Chlamydia trachomatis [14]. In this study, only one participant reported having chlamydia, but no positive results were found. It is worth noting that the study’s objective did not include the analysis of co-infections, which are known to be highly relevant.

In Colombia, the diagnostic algorithm for HPV provided by the Ministry of Health and Social Protection proposes diagnostic strategies by age group and includes screening between ages 25 and 65. It establishes different strategies such as combined HPV and cytology screening every five years or exclusive Pap smear every three years [21,22].

The low sensitivity of cytology has necessitated the inclusion of HPV detection platforms using molecular biology techniques as additional or initial diagnostics and very promising results have been obtained, particularly with combined techniques [8]. This type of molecular testing for HPV detection and genotyping has significant diagnostic value in cervical cancer surveillance and prevention programs, as it allows for the selection of women with histological lesions that may progress to high-grade lesions or carcinoma, thus reducing the costs associated with cervical cancer diagnosis [13].

This study evaluated the prevalence and genotypes of HPV in cervicovaginal and urine samples, finding a general HPV prevalence of 29.2% and 32.9%, respectively, in contrast with studies reporting 68.8% and 62.9% positivity in cervicovaginal and urine samples [23]. In Colombia, a prevalence of 60% and 64.7% has been reported [24]. However, the figures in this study are higher than those found by Dong *et al*. [25]. These differences could be attributed to the type of sample evaluated, the distribution of the virus, and the viral load at the time of sampling [26].

When comparing cervicovaginal and urine samples, a weak concordance (kappa=0.392) was observed, with a sensitivity of 61.7% and specificity of 78.9% for viral detection in urine. These findings contrast with other studies that have reported moderate to almost perfect concordance between the two sample types [24,25,27]. However, the results suggest that, under the conditions of this study, urine samples are not an adequate routine method for HPV detection.

Regarding genotyping, high-risk genotypes were identified in both sample types, with the most prevalent being types 52 and 68. These results differ from other studies in Colombia and Latin America, where higher prevalence of genotypes 16 and 18 have been reported [7,28,29]. This variability in genotype distribution highlights the importance of early genotyping in the clinical context and cervical cancer prevention programs. While persistent infection with oncogenic HPV genotypes is necessary for cervical carcinogenesis, other poorly defined molecular factors and events drive the malignant transformation of the cervical epithelium over a long period of infection [17].

This study provides valuable information on HPV prevalence and genotyping in a population of women from Tunja, Colombia. Although high-risk genotypes were identified, the low concordance and diagnostic performance of viral detection in urine highlight the need to improve detection techniques for these types of samples for potential implementation as a non-invasive screening method.

## 5. CONCLUSIONS

The prevalence of HPV infection in women residing in Tunja is lower compared to national and international studies, with high-risk genotypes present in both cervicovaginal and urine samples. The diagnostic performance of the urine test is low, making it unsuitable for diagnosing infection and determining genotypes due to the heterogeneity of results between the two samples.

## Data Availability

All relevant data are included in the manuscript and its supporting information files.

## 6. ACKNOWLEDGMENTS

The authors thank the women who participated in the research.

## Contributors

Lorenzo Hernando Salamanca Neita: study conception and design, population invitation, survey administration, article writing, critical review of the document, and final approval of the version to be presented.

Elizabeth Guío Mahecha: study conception and design, article writing, critical review of the document, and final approval of the version to be presented.

Mónica Gabriela Huertas: study conception and design, article writing, critical review of the document, and final approval of the version to be presented.

Johana Marín Suárez: study conception and design, sample processing, data analysis and interpretation, article writing, critical review of the document, and final approval of the version to be presented.

Juan Pablo Carvajal Rojas: study conception and design, article writing, critical review of the document, and final approval of the version to be presented.

Gloria Eugenia Camargo Villalba: study conception and design, population invitation, sample collection, data analysis and interpretation, article writing, critical review of the document, and final approval of the version to be presented.

Laura Ximena Ramírez López: study conception and design, population invitation, survey administration, data analysis and interpretation, article writing, critical review of the document, and final approval of the version to be presented.

